# Dynamic functional synchronization profiles in autism differ by spatial scale and along hierarchical cortical gradients

**DOI:** 10.64898/2025.12.11.25342053

**Authors:** Benedikt P. Fuhr, Yonatan Sanz Perl, Ines Severino, Morten L. Kringelbach, Gustavo Deco, Henricus G. Ruhé, Michael V. Lombardo

**Affiliations:** Laboratory for Autism and Neurodevelopmental Disorders, Center for Neuroscience and Cognitive Systems, Istituto Italiano di Tecnologia, Rovereto, Italy; Center for Mind/Brain Sciences, University of Trento, Rovereto, Italy; Center for Brain and Cognition, Computational Neuroscience Group, Faculty of Medicine and Life Sciences, Universitat Pompeu Fabra, Roc Boronat 138, Barcelona, 08018, Spain; Institució Catalana de la Recerca i Estudis Avançats (ICREA), Passeig Lluís Companys 23, Barcelona, 08010, Spain; Centre for Eudaimonia and Human Flourishing, Linacre College, University of Oxford, Oxford, UK; International Centre for Flourishing, Universities of Oxford (UK), Aarhus (Denmark) and Pompeu Fabra (Spain); Department of Clinical Medicine, Aarhus University, Aarhus, Denmark; Department of Psychiatry, University of Oxford, Oxford, UK; Department of Psychiatry, Radboudumc, Nijmegen, Netherlands; Donders Institute for Brain, Cognition and Behavior, Radboud University, Nijmegen, Netherlands

## Abstract

Although prevailing theories propose that autism is characterized by local cortical overconnectivity and long-range underconnectivity, the supporting empirical evidence has been mixed. Here we demonstrate that these divergent findings can be reconciled by examining how functional synchronization profiles dynamically change over time and across the cortex over different spatial scales. We applied the turbulence dynamics framework to the ABIDE dataset (n = 1,009) and found evidence for altered functional synchronization dynamics in autism. Autistic individuals showed increased short-range and reduced long-range functional synchronization variability over time, as well as reduced synchronization strength across all spatial scales. Synchronization also decayed more rapidly with distance and exerted weaker influence across scales in autism. These distance-specific alterations suggest that local hyperconnectivity may generate turbulent, chaotic synchronization dynamics that fail to propagate coherently across the cortex, resulting in an overly rigid brain organization at longer distances. Mapping these effects onto the sensorimotor–association cortical gradient revealed increased variability in sensorimotor regions and decreased variability in the association cortex. Together, we found evidence of disturbances in functional synchronization dynamics at different spatial scales and along hierarchical brain gradients in individuals with autism. These results help consolidate ideas about how dynamic functional connectomic organization manifests in autism and pinpoints likely early neurodevelopmental effects along a primary axis of hierarchical cortical organization.

## Introduction

Atypical neural connectivity has long been proposed to be a convergent neurobiological explanation behind the many ‘autisms’ (Belmonte et al., 2004; Courchesne & Pierce, 2005; Geschwind & Levitt, 2007). Such atypical neural connectivity has been shown to manifest in multiple ways ranging from genetic alterations to specific key synaptic proteins, convergent gene expression dysregulation affecting synaptic processes, to observations of differences in macroscale structural and functional brain connectivity measured from neuroimaging data (e.g., Bourgeron, 2015; De Rubeis et al., 2014; Di Martino et al., 2014; Gandal et al., 2022; Holiga et al., 2019; Pagani et al., 2025). Several types of intrinsic functional connectivity differences measured from resting state fMRI (rsfMRI) data have been observed throughout the literature. For example, based on static functional connectivity analysis (i.e. analysis of the entire rsfMRI time-series), autism can be characterized by alterations in whole-brain functional connectivity gradients (Hong et al., 2019), idiosyncratic connectivity profiles (Benkarim et al., 2021; Hahamy et al., 2015; Nunes, 2019), mosaic patterns of hypo- and hyper-connectivity (Di Martino et al., 2014; Holiga et al., 2019; Ilioska et al., 2023; Pagani et al., 2025), as well as distance-dependent effects (Courchesne & Pierce, 2005; Weber et al., 2024).

While static functional connectivity captures the average connectivity pattern over an entire scan, they assume connectivity patterns are somewhat stationary over time. Alternatively, other studies have demonstrated that rsfMRI showcases intrinsic non-stationarities or ‘dynamics’ (Calhoun et al., 2014) that necessitate approaches that analyze how dynamic functional connectivity patterns emerge and change over time (Preti et al., 2017). Within autism, dynamic functional connectivity approaches have suggested potentially hyper-variant and weaker functional dynamics (Chen et al., 2017; Mash et al., 2019). However, findings across the literature remain mixed (Hull et al., 2017; Mohammad-Rezazadeh et al., 2016).

While dynamic functional connectivity investigates changes of statistical dependencies between different parts of the brain over time, it primarily characterizes *when* and *how* changes in the data pattern occur, rather than *why* they occur. To move beyond description and toward a mechanistic understanding of these dynamic processes, recent work has emphasized the role of *metastability* - a dynamical systems concept that captures the brain’s ability for flexible switching between different functional configurations (Deco et al., 2017; Hancock et al., 2025; Tognoli & Kelso, 2014). Alterations in metastable dynamics have been used to distinguish between different psychiatric disorders, such as Alzheimer’s disease (Alderson et al., 2018) and schizophrenia (Hancock et al., 2023). In autism, metastability has been shown to offer a promising framework to link neuronal and behavioral flexibility. Core features like restricted repetitive behavior and associated behavioral inflexibility have been linked to impaired neural state transitions (Watanabe & Rees, 2017), and recent evidence suggests that modifying this neuronal inflexibility can directly enhance behavioral flexibility (Watanabe & Yamasue, 2025).

However, metastability is typically used to characterise the configuration of the *whole* brain. Concepts from other disciplines, such as fluid dynamics, can extend metastability to capture spatial differences more effectively. One such framework is *turbulence*, an approach that draws on principles from fluid dynamics, a branch of physics. Flow can be classified as either laminar or turbulent. Laminar flow is characterised by the smooth, orderly movement of fluid particles in parallel layers, resulting in predictable behavior. In contrast, turbulent flow is characterized by chaotic and irregular motion. The formation of vortices of different spatial scales makes turbulent flow highly unpredictable (Kalsi & Balani, 2016). An important property of turbulent flow is its ability to rapidly transfer energy across spatial scales, which allows information to propagate through the system more efficiently (Kolmogorov, 1991). Deco and Kringelbach (2020) combined these properties with the theoretical framework of Yoshiki Kuramoto (1984), who generalized turbulence beyond the limited domain of fluid dynamics to other physical systems including coupled oscillators (Deco et al., 2025). Deco and Kringelbach have long worked on whole-brain modelling of brain dynamics and shown that one of the best models of the empirical data uses coupled Stuart-Landau Oscillators (Deco & Kringelbach, 2025). Their research models the brain as a system of coupled oscillators that can help to identify turbulent markers in the brain with rsfMRI. This demonstrated the value of the framework in providing a more comprehensive and fine-grained analysis of spatiotemporal synchronization dynamics (Deco & Kringelbach, 2020).

This work presents an investigation into the dynamics of functional synchronization across different spatial scales in rsfMRI in autism. We tested the hypothesis that these dynamics differ between individuals with autism and those with typical development (TD). First, we hypothesize that changes in local synchronization, and consequently, local turbulence values are increased in autism due to local hyperconnectivity. At a more global scale, however, we hypothesize that turbulence values are reduced in people with autism compared to TD individuals. Second, we hypothesize that synchronization cascades and synchronization transfer differ across and within spatial scales in autism. To test these hypotheses, we applied the turbulence dynamics framework to rsfMRI scans from the Autism Brain Imaging Data Exchange (ABIDE) datasets.

## Methods

### Dataset

For the present study, we used the Autism Brain Imaging Data Exchange repositories (ABIDE I and II, Di Martino et al., 2014, 2017). ABIDE I consists of a total of n = 1,112 participants (n = 539 of whom have been diagnosed with Autism) from 17 different sites, whereas ABIDE II consists of n = 1,114 participants (n = 521 of whom have been diagnosed with Autism) from 19 different sites. From the total ABIDE I and II datasets, we included data from 24 sites. Data from the University of Michigan was excluded from the analysis due to its imbalanced distribution of Autism and TD (∼6:1 Autism to TD).

We implemented further quality checks and filtering criteria after preprocessing of the resting state scans. Subjects which demonstrated poor image quality following visual inspection (e.g., failed spatial normalization or co-registration) were excluded from the study. Furthermore, subjects with a mean framewise displacement (FD) above 0.5mm were excluded from the study. In order to minimize confounds of motion on our analysis, we undertook a further investigation of group differences of mean FD between autism and TD. A statistically significant increase in movement was observed in the Autism group in comparison to the TD group (*t*(1157) = 4.17, *p* = 0.00003, *d* = 0.26). To address this issue, a matching process was performed on both groups according to their FD-values. This was achieved by employing the *matchit* function from R, with a caliper of 0.05 (Ho et al., 2011). Following the implementation of this matching process, the differences observed between the groups in terms of FD were found to be non-significant (*t*(1010) = 0.6165, *p* = 0.538, *d* = 0.039). After applying these filtering criteria, a total of n = 505 autistic individuals (mean age = 16.29 years, age SD = 9.00) and n = 504 non-autistic individuals (mean age = 16.68 years, age SD = 8.94, Supplementary Table S1) remained.

### Imaging parameters and fMRI preprocessing

The preprocessing of the resting state scans follows the pipeline described in Trakoshis et al., 2020. Preprocessing of the resting state data was split into two components: core preprocessing and denoising. Core preprocessing was implemented with AFNI (Cox, 1996) using the tool speedypp.py (Kundu et al., 2012). This core preprocessing pipeline included the following steps: (i) slice acquisition correction using heptic (7th order) Lagrange polynomial interpolation; (ii) rigid-body head movement correction to the first frame of data, using quintic (5th order) polynomial interpolation to estimate the realignment parameters (three displacements and three rotations); (iii) obliquity transform to the structural image; (iv) affine co-registration to the skull-stripped structural image using a gray matter mask; (v) nonlinear warping to MNI space (MNI152 template) with AFNI 3dQwarp; (vi) spatial smoothing (6 mm FWHM); and (vii) a within-run intensity normalization to a whole-brain median of 1000. Core preprocessing was followed by denoising steps to further remove motion-related and other artifacts. Denoising steps included: (viii) wavelet time series despiking (‘wavelet denoising’); (ix) confound signal regression including the six motion parameters estimated in (ii), their first order temporal derivatives, and ventricular cerebrospinal fluid (CSF) signal (referred to as 13-parameter regression). The wavelet denoising method has been shown to mitigate substantial spatial and temporal heterogeneity in motion-related artifacts that manifest linearly or nonlinearly and can do so without the need for data scrubbing (Patel et al., 2014). Data scrubbing (i.e. volume censoring) cannot be used in our time-series-based analyses here as such a procedure breaks up the temporal structure of the time-series. Wavelet denoising is implemented with the Brain Wavelet toolbox. The 13-parameter regression of motion and CSF signals was achieved using AFNI 3dBandpass with the –ort argument.

The ABIDE dataset varies in terms of scanner types and other types of imaging parameters (see details of imaging parameters by site in Supplementary Table S2). Since these types of variation between imaging sites can affect BOLD data and connectivity measurements, we implemented a site correction technique such as ComBat (Leek et al., 2012). This has been applied after the main preprocessing pipeline, but before any tests of group differences were made. See the section on Statistical Analysis for more details.

### Turbulence analysis

Average BOLD time series data was extracted from a custom-made parcellation scheme, which combined the 7 network Schaefer-1000 atlas (Schaefer et al., 2018) and the Melbourne subcortical atlas (Tian et al., 2020). This custom-made atlas comprised 1,054 regions, 54 of which were subcortical. After extraction of average BOLD time series data for each parcel, data was then transformed to phase-space using the Hilbert transformation. Consequently, we obtained instantaneous phase information for each of the 1,054 parcels as a function of time, which served as the basic input matrix for all subsequent analyses.

This study applies the turbulence dynamics framework, which has recently been employed to analyze dynamic synchronization patterns in human brain activity, (Deco & Kringelbach, 2020, see Figure 1). To formalize these turbulent dynamics, we adopt the approach of viewing the brain as a complex system of coupled oscillators (Kuramoto, 1984). In this model, the time-series of the 1,054 brain parcels serve as fundamental elements of the system. Information for each parcel is provided by the instantaneous phase of the Hilbert-transformed time series (Figure 1A and 1B). To quantify the magnitude of synchronization at a given time point, we computed the Kuramoto order parameter, denoted by the letter R. This order parameter was computed either uniformly over the whole brain (Kuramoto *global* order parameter, henceforth referred to as ’KGOP’), or over a range of spatial scales (denoted by the coefficient λ), which resulted in the Kuramoto *local* order parameter (henceforth referred to as ’KLOP’) (see Figure 1C–E). KGOP values indicate the extent to which the entire brain is in phase synchrony at a given time. In contrast, KLOP values indicate the degree to which a given parcel is in phase synchrony with a selected portion of its neighbouring regions at a given time. The ’selection’ of these neighbouring regions is determined by the spatial scale λ, which ultimately acts as a kernel function for the given brain parcel. Here, λ ranges from 0.01 to 0.28 in ten steps of 0.03 and indicates the extent to which neighbouring regions contribute to the level of synchronization (Figure 1D). A higher value of λ (e.g., 0.28) corresponds to a more constricted weighing kernel, where influence diminishes more rapidly with increasing distance. Consequently, regions further away contribute less to the KLOP of the specified brain parcel. Conversely, a lower value of λ (e.g., 0.01) corresponds to a wider weighing kernel. Therefore, the influence diminishes more gradually with increasing distance, and regions that are further away contribute more to the KLOP of the specified brain area (Figure 1E, Supplementary Movie S1).

**Figure 1.**
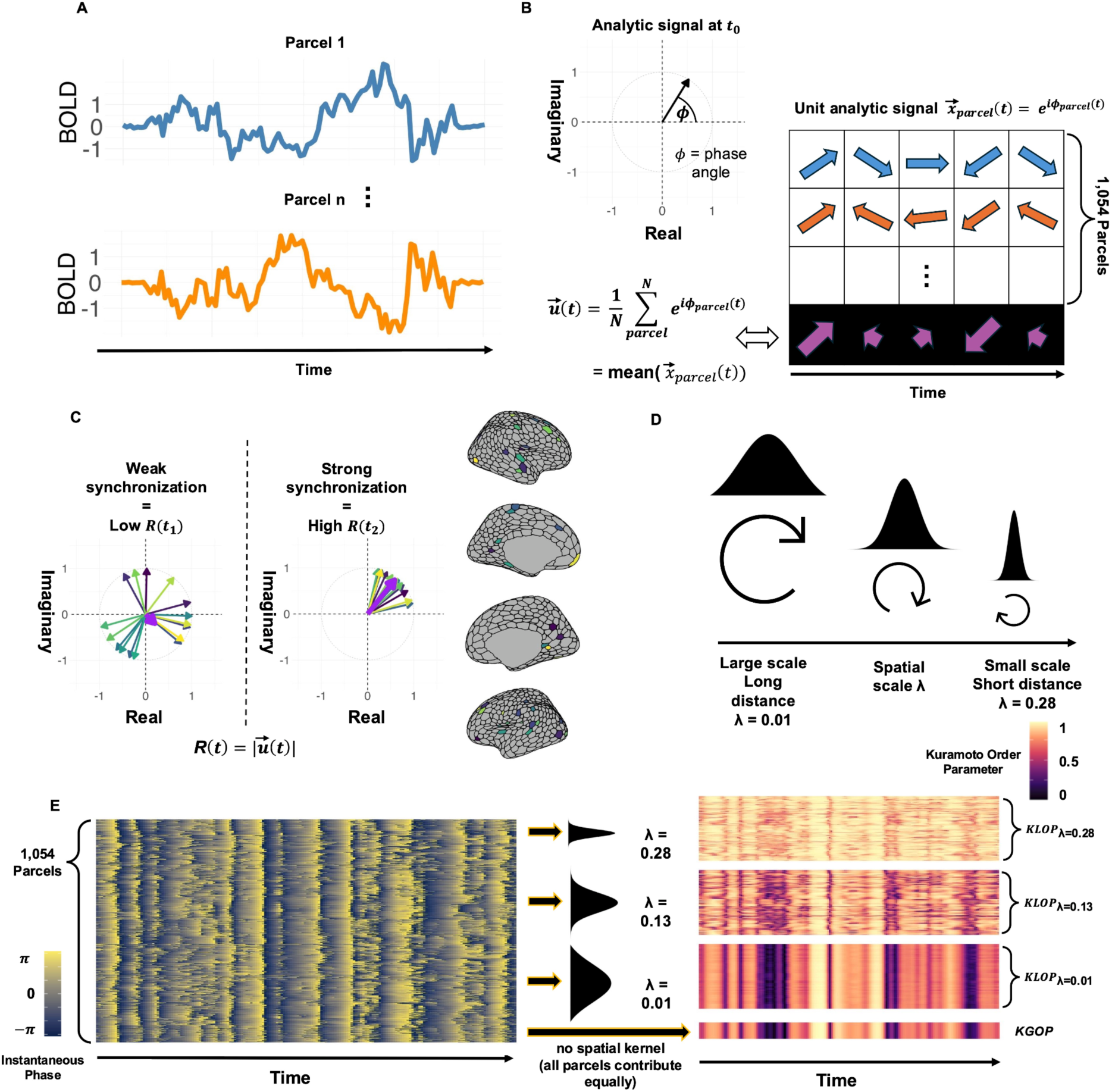
Methodology and turbulence dynamics framework. (A) The BOLD time series for 1,054 parcels was extracted using a combined 1,000-parcel cortical atlas (Schaefer et al., 2018) and a 54-parcel subcortical atlas (Tian et al., 2020). (B) The BOLD time series for each parcel was transformed using the Hilbert transform to obtain the analytic signal. For a given parcel, each time point in the time series then contains two-dimensional information that can be represented as a vector (top left panel). Each vector is composed of two parts: a real part (Re, x-axis), containing the value of the original BOLD time series, and an imaginary part (Im, y-axis), containing the 90° phase-shifted version of that value. Using Euler’s formula, this vector is expressed in terms of instantaneous phase (angle ϕ) and amplitude. Since our focus is on synchronization, we extracted only the phase information by normalizing each vector to unit length, resulting in the complex unit analytic signal x(t) (right panel). At each time point, the unit analytical signal for each parcel (illustratively shown for two parcels in blue and orange) was summed and then divided by the total number of parcels to obtain the mean unit vector u(t) (purple arrows at the bottom). (C) The Kuramoto order parameter R is defined as the magnitude (or length) of u(t) and varies as a function of time. The two panels show a graphical representation of R at two different time points, t₁ and t₂. Each colored vector resulting from the Hilbert-transformed time series represents the unit analytic signal x(t) of a parcel of the Schaefer-Tian parcellation. The location of each parcel in the brain can be seen in the plots on the right. The bold purple vector represents the mean unit vector; its length is R. When the colored vectors are dispersed, they cancel each other out. The resulting sum is relatively small, leading to a small R (left-hand panel, weak synchronization). When the vectors are aligned in phase, the resulting summed vector is relatively long. This leads to a higher R (right-hand panel, strong synchronization). (D) Spatial scales were introduced to investigate synchronization and information transmission across different distances in the brain. Low values of λ correspond to larger spatial scales/longer distances in the brain. Conversely, as λ increases, the spatial scale decreases, and synchronization at more local levels is investigated. (E) To compute a localized version of the Kuramoto order parameter for a single parcel n₀, the instantaneous phase of all other parcels (left panel) is combined with a spatial kernel function (middle panel) that weighs their contributions based on their Euclidean distance to n₀. For each parcel n and each kernel width λ, the weighted unit analytic signal is summed, following the standard Kuramoto formula (see Figure 1B). The magnitude of the resulting complex average yields the Kuramoto local order parameter (KLOP), which varies as a function of time. The rightmost panel shows two-dimensional heatmaps of the KLOP for three different values of λ. Each heatmap shows how the KLOP changes over time, with the rows representing different parcels of the brain and the columns representing successive time points. Higher KLOP values, indicating stronger synchronization, are shown in brighter colors, while lower values (weaker synchronization) appear darker. The λ decreases (i.e. the spatial scale increases) from the topmost to the bottommost heatmap. This results in a gradual loss of temporal and spatial detail. It reflects a shift from a more focal kernel capturing local synchronization (top heatmap) to a broader kernel assessing more global synchronization (bottom heatmaps). To contrast the KGOP with the KLOP, the KGOP is visualized as a one-dimensional vector at the very bottom. The KGOP is computed without any weighting kernel and therefore captures the phase synchronization of the whole brain, where each parcel contributes equally to the magnitude of synchronization. Note that, even though some spatial detail is preserved, the KLOP approximates the KGOP for each parcel on very large spatial scales (i.e. λ = 0.01).

To summarize, the KLOP formalizes the turbulence dynamics framework by quantifying *how much* (the scalar value) and *over which distance* (the spatial scale) synchronization is exerted in the brain. The three-dimensional KLOP matrices, where the rows represent parcels and the columns represent timepoints, and the third dimension represents the λ values, were used in the subsequent analysis to compute several higher-order statistics. The methodologies employed to extract these are described in detail in Escrichs et al. (2022) and Martínez-Molina et al. (2024). For a detailed exposition of the initial whole-brain dynamics framework employing turbulence, reference should be made to Deco and Kringelbach (2020). The computation and extraction of these measures were implemented in MATLAB version 2024b. Here we outline the rationale behind each of these higher order statistics.

First, we computed four higher-order statistics using the *Kuramoto Local Order Parameter* (KLOP) for each λ, making them susceptible to spatial scales:

**(1) Amplitude turbulence** captures the spatio-temporal extension of metastability. For each spatial scale, a slice of the three-dimensional KLOP matrix is vectorized, and the standard deviation is computed on the resulting vector. This is therefore a measure of the variability of synchronization on a given spatial scale (Figure 2a).
**(2) Scale-specific synchronization** captures the strength of synchronization at a given distance. For each spatial scale, a slice of the three-dimensional KLOP matrix is vectorized, and the mean is computed on the resulting vector. This is therefore a measure of the central tendency of synchronization at a given spatial scale (Figure 2a).
**(3) Information cascade flow** captures the propagation of synchronization from a lower spatial scale to its immediately adjacent higher spatial scale. This is calculated by advancing the KLOP values of a given parcel’s higher spatial scale by one timestep (lower λ-value) and correlating the resulting vector with the parcel’s original KLOP vector at the adjacent lower spatial scale (higher λ-value). This correlation value is computed for each parcel and then averaged. It measures the extent to which synchronization at the lower spatial scale influences, or is reflected in, synchronization at the higher spatial scale for each pair of adjacent spatial scales (Figure 2b).
**(4) Information transfer** captures the spatial extent over which synchronization is maintained within the brain at a given spatial scale. It is quantified by correlating the KLOP values across all pairs of parcels within that scale. These correlation values are then modeled as a function of inter-parcel distance, using a linear fit. The slope of this fit reflects the rate at which synchronization strength decays with distance (Figure 2c).

**Figure 2.**
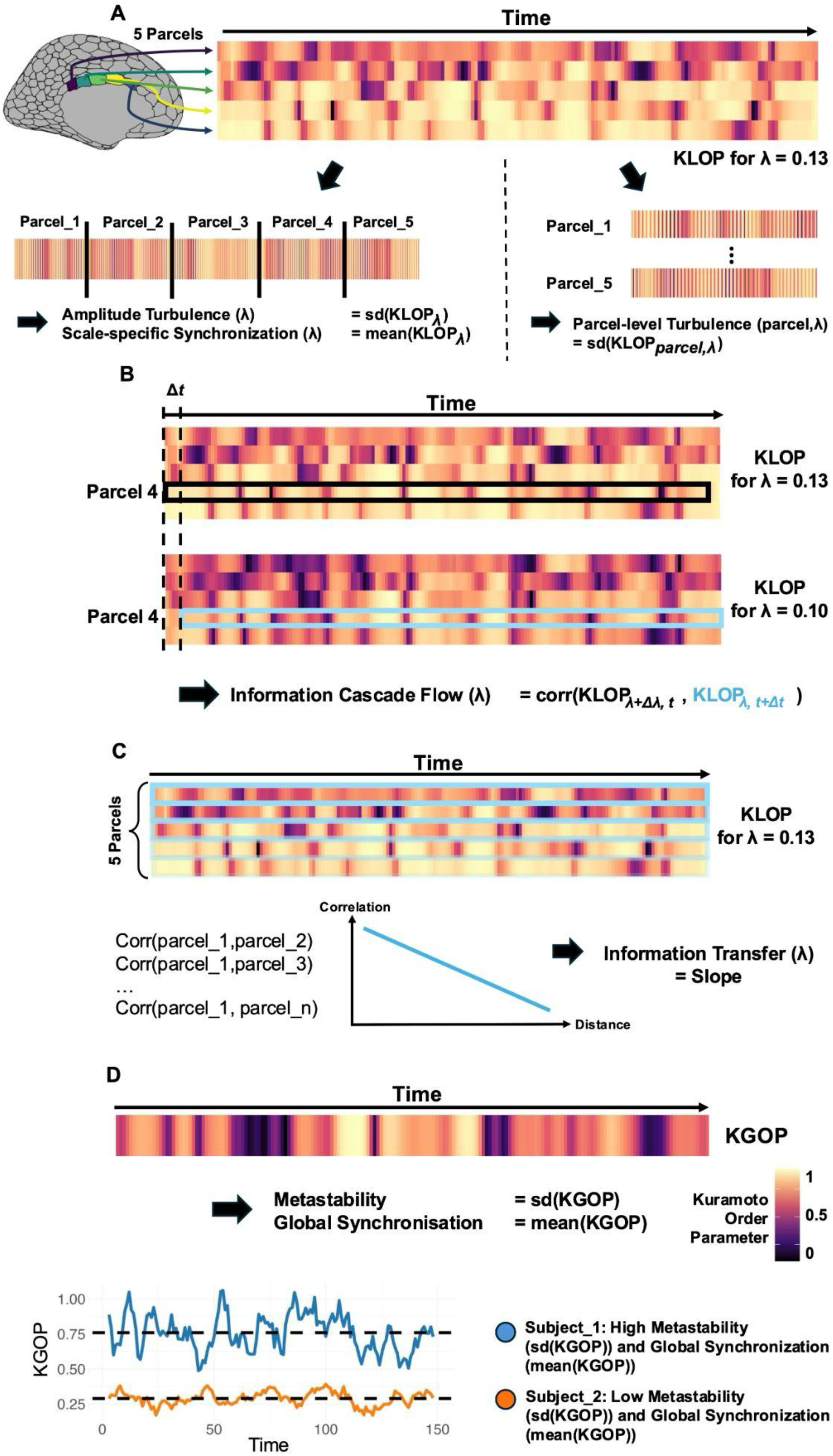
Description of higher-order and whole-brain summary statistics. Panel A illustrates the computation of amplitude turbulence, parcel-level turbulence and scale-specific synchronization statistics. The heatmap at the top shows the evolution of the Kuramoto Local Order Parameter (KLOP) for a given spatial scale (λ=0.13) for five representative parcels over time. Each row represents a parcel, with the distance between parcels increasing from top to bottom. Each column represents a time point, with the color scale representing KLOP values. To compute the scale-specific synchronization and amplitude turbulence for a given spatial scale, this two-dimensional matrix is vectorised (bottom left of the panel). The standard deviation of the resulting one-dimensional vector over time is the amplitude turbulence for this spatial scale, while the mean is the scale-specific synchronization. The bottom right of the panel shows how parcel-level turbulence is computed for each parcel and spatial scale. The standard deviation of a parcel’s KLOP for a given λ is that parcel’s turbulence. Panel B visualizes the computation of the information cascade flow. The two heatmaps show the evolution of the Kuramoto Local Order Parameter (KLOP) for two spatial scales (λ=0.13 and λ=0.10) for five representative parcels over time. The KLOP for parcel 4 is highlighted with a box. The information cascade flow is calculated by advancing the KLOP values of parcel’s 4 higher spatial scale (λ=0.10, blue box, bottom heatmap) by one timestep to the right and correlating the resulting vector with the parcel’s 4 original KLOP vector at the adjacent lower spatial scale (λ=0.13, black box, top heatmap). This correlation value is computed for each parcel, and then averaged to obtain the information cascade flow for this λ pair. Panel C visualizes the computation of the information transfer statistics. For a given spatial scale (e.g., λ=0.13), the correlation between each parcel and all other parcels is computed. The resulting correlation values are fitted with a line as a function of distance. The slope of this linear fit then represents the information transfer value for this spatial scale. Panel D shows the computation of whole-brain summary statistics for metastability and global synchronization. The one-dimensional vector is the Global Kuramoto Order Parameter (KGOP). Metastability statistics are the standard deviation of the KGOP over time, and global synchronization statistics are the mean of the KGOP over time. Metastability and Global Synchronization for two hypothetical subjects are visualized at the very bottom.

Second, we computed two whole-brain summary statistics using the *Kuramoto Global Order Parameter* (KGOP). The KGOP is computed using the same rationale as the KLOP. Unlike the KLOP, however, the KGOP captures the synchronization of the whole brain at a given moment in time, instead of the synchronization for each individual parcel. Consequently, it does not use a Gaussian weighting kernel in its computations, so all parcels have the same weight. The KGOP is then computed as the magnitude of the mean unit vector u(t) (see Figure 1b and 1c). Note, however, that for the lowest λ values (e.g., λ = 0.01, indicating very large spatial kernels), the KLOP approaches the KGOP since the kernel function becomes wider and all parcels therefore contribute almost equally to the Kuramoto order parameter (note the similarity between the KLOP (λ = 0.01) and KGOP heatmaps at the bottom of Figure 1e). We computed two summary-statistics:

**(5) Metastability** is a heuristic measure of the configuration of the attractor landscape underlying complex brain dynamics and captures the temporal variation in whole-brain synchronization (Hancock et al., 2025). Here, it is computed as the standard deviation of the KGOP over time (Figure 2d).
**(6) Global synchronization** captures the central tendency of the whole-brain synchronization and is computed as the mean of the KGOP over time (Figure 2d).

Third, we computed parcel-level turbulence for different values of λ. For each λ, parcel-level turbulence is calculated using the standard deviation of a given parcel’s KLOP vector over time (Figure 2a). This captures how a parcel’s synchronization varies over time at different spatial scales.

### Statistical analysis

After fMRI preprocessing, quality control, and turbulence analysis pipelines were implemented, we addressed the issue of variables that differ between sites with a site correction technique known as *ComBat* (Leek et al., 2012). Implementation of this analysis was achieved using the ComBat function within the *sva* R library. The ComBat model included variables such as diagnosis, sex, age, mean FD and a diagnosis×age interaction as covariates in the model. To examine the effect of ComBat for removing variance due to site, we compared the first principal component from a PCA analysis before and after implementing ComBat. PC1 before ComBat implementation shows substantial variability between sites (*F(23,985)* = 11.99, *p* < 0.001, Figure S1, Panel A). This variability is eliminated after ComBat implementation, which demonstrates the effectiveness of ComBat at removing site variation (*F(23,985)* = 1.053, *p* = 0.394, Figure S1, Panel B).

To assess group-difference in whole-brain summary measures (i.e., metastability and global synchronization), we used linear models implemented with the *lm* function in R. The model included sex, mean FD, diagnosis, age, and the diagnosis×age interaction as independent variables. The dependent variable was one of metastability or global synchronization. To assess group-difference over measures using different spatial scales (i.e., amplitude turbulence, scale-specific synchronization, information cascade flow and information transfer) we used linear mixed effect models, implemented with the *lmer* function in the *lme4* R library. Fixed effects included diagnosis, λ, age, sex, and mean FD, along with interaction terms for diagnosis×λ and diagnosis×age. Random effects were modeled for the repeated measure of λ with random intercepts and slopes. Post-hoc pairwise comparisons were used to compare groups at each value of λ by computing the estimated marginal mean (EMM) from the model and performing pairwise *t*-tests for each spatial scale. To account for multiple comparisons, p-values from the post-hoc tests were corrected using the False Discovery Rate (FDR) method.

To assess differences in parcel-level turbulence between TD and autism across λ levels, we calculated Cohen’s d as the difference between the two groups for each of the 1,054 parcels. We then extracted the 100 parcels that showed the biggest absolute value difference in parcel-level turbulence for each λ. For each spatial scale, these 100 parcels were attributed to one of seven RSNs (Yeo et al., 2011) and a subcortical network to determine which networks are enriched for the biggest differences in parcel-level turbulence.

Discrete RSNs demonstrate that the brain is intrinsically functionally organized. However, to provide a different, yet complementary perspective to RSNs, we also examine continuous gradients as another key principle for explaining how the brain is hierarchically organized. Thus, in addition to looking at turbulence group differences relative to known RSNs, we also examined how similar turbulence group differences would be with continuous gradients of hierarchical cortical organization, such as the sensorimotor-association (S-A) axis (Sydnor et al., 2021). The S-A axis is a multimodal hierarchical gradient that describes how many features (e.g., macrostructural, microstructural, functional, metabolic, transcriptomic, and evolutionary) vary across the cortex (Sydnor et al., 2021). As an example, it is well known that the S-A axis is quite similar in topography to the primary functional connectivity (FC) gradients measured with rsfMRI (Margulies et al., 2016). The S-A axis is also neurodevelopmentally sensitive (Sydnor et al., 2021; Tsyporin et al., 2025) and thus allows an interesting perspective on neurodevelopmental disorders like autism. One pole of the S-A or FC axes comprises areas involved in sensorimotor and unimodal processing, while the other pole consists of association areas and transmodal processing areas. The S-A axis is hypothesized to be a neurodevelopmentally important axis for hierarchical brain organization in autism (Bernhardt et al., 2025) and may be one of the downstream convergent consequences of early genomic cortical patterning differences in autism (Gandal et al., 2022; Lombardo et al., 2021). In these analyses, we evaluate the similarity between parcel-level turbulence group difference maps and S-A and FC gradient maps. To achieve this we first identified the top 100 parcels with the strongest group differences in parcel-level turbulence. Ranking parcels to get a top 100 involved first computing a rank score for all 1,000 cortical parcels from the Schaefer parcellation. We then computed a weighted sum of the Sydnor gradient rank score from the five nearest neighbours for each of the 1,000 parcels, with the weight dependent on the distance. Then, we selected the top 100 parcels showing biggest differences in effect sizes for each λ and assigned a gradient score to each of the 100 parcels. We then fitted linear, polynomial, and logistic models to best characterize how the gradient score of the top 100 parcels (i.e the dependent variable) could be predicted from λ values (i.e. the independent variable). We then performed model selection between these three models and utilized a combination of the residual sum of squares (RSS), Akaike information criterion (AIC) and Bayesian information criterion (BIC) criteria to identify the best fitting model. Once the best model has been identified, we then report inferential statistics to describe how the gradient may relate to effect sizes over spatial scales. In addition to examining the top 100 parcels, we also adopted a different approach of simply testing for correlations between parcel-level turbulence group difference maps and S-A and FC maps. For this analysis, we used the same Schaefer 400 parcellation as Sydnor et al. (2021) and computed parcel-level turbulence values for each of the 400 parcels by taking the weighted average of the five closest parcels from the initial parcellation of 1,000 cortical parcels. Standardized effect size (Cohen’s d) was then computed for every parcel and this was repeated at each level of λ. Statistical comparisons were then made between parcel-level Cohen’s d turbulence maps and the S-A or FC maps. These were implemented by computing Spearman ρ and then evaluating statistical significance via permutation p-values from ‘spin tests’ (10,000 repetitions) that preserve spatial autocorrelation (Alexander-Bloch et al., 2018; Markello & Misic, 2021). To account for multiple comparisons, we corrected the p-values from the permutation tests using the false discovery rate (FDR) method.

All code to reproduce the analyses presented in this paper is available under https://gitlab.iit.it/bmp006-public/LAND_IIT/turbulence_abide/.

## Results

### Autism is characterized by higher variability in synchronization at local/small spatial scales

This study applied the turbulence-dynamics framework to rsfMRI data from the ABIDE dataset (autism n = 505; TD n = 504). First, we examined differences in synchronization variability by looking at our whole-brain measure of synchronization (KGOP) using a metric called metastability (i.e., sd(KGOP)). Metastability was significantly different between-groups (see Figure 3a and Supplementary Table S3, *F*(1,1003) = 3.92, *p* = 0.048, *d* = 0.13), with lower levels in the autism group than in the TD group. This result indicates that whole-brain synchronization is less variable over time in autism. While this initial analysis assessed variability of functional synchronization at the largest spatial scale (i.e., the whole brain), our framework also allows us to examine synchronization at finer spatial scales, potentially revealing group differences at more localized levels. Thus, in our next analysis, we used a distance-specific weighting parameter, denoted as λ, to assess group differences across different spatial scales. For our analyses, λ is inversely linked to spatial distance; smaller values of λ indicate larger spatial scales, and higher values of λ represent smaller, more localized spatial scales. We then used the resulting distance-specific measures of synchronization (i.e., KLOP) to examine how functional synchronization variability (i.e., sd(KLOP) or ‘*amplitude turbulence*’) changes with spatial scale (λ). Although we did not find a main effect of diagnosis on amplitude turbulence levels (*F*(1, 1041) = 0.09, *p* = 0.763), we did find a significant λ×diagnosis interaction (*F*(1, 1198) = 8.58, *p* = 0.003; see Figure 3b and Supplementary Table S4). Follow-up case-control comparisons at each level of λ revealed significantly higher variability in functional synchronization for autism at smaller or more localized spatial scales. However, this group difference was eliminated as spatial scale increased (i.e. decreased λ). This suggests that compared to TD, synchronization over shorter distances varies more over time in autism. However, as the spatial distance increases, temporal fluctuations in synchronization normalize and are not different between TD and autism. This result of higher variability in functional synchronization at smaller or more localized spatial levels in autism is striking given that at the whole brain level (see Figure 3a), the difference is reversed in autism (e.g., less variability in functional synchronization in autism). This strong contrast is important for validating the idea that functional synchronization is more turbulent at small, localized spatial scales and less turbulent at the whole-brain level in autism.

**Figure 3.**
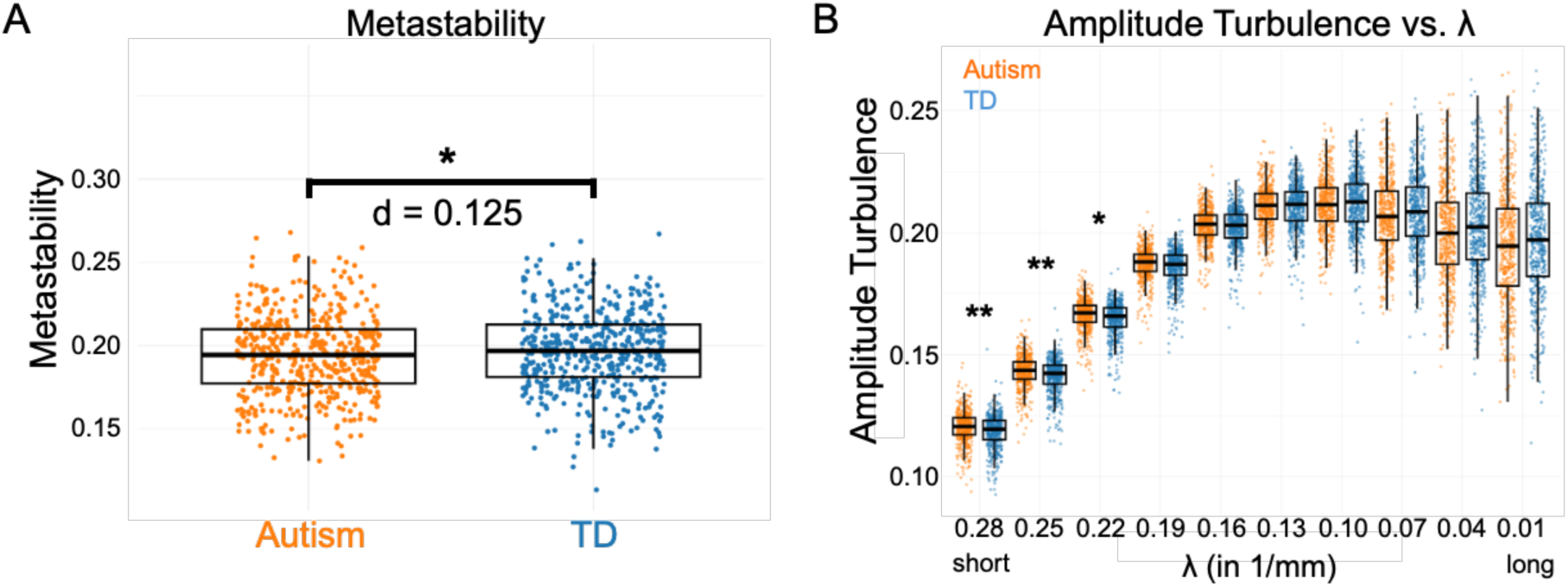
Increased synchronization variability at small spatial scales in autism. Panel A shows a significant effect of a group difference between autism and TD in whole-brain variability of synchronization (e.g., metastability). Autism shows a small effect of lower levels of metastability at the level of the whole-brain relative to TD. (B) Amplitude turbulence levels of autism (orange) vs. TD (blue) as a function of spatial scale (λ). Amplitude turbulence levels are significantly increased in autism relative to TD for smaller spatial scales (i.e. high λ = shorter distances). As the spatial scale increases (i.e. larger distances), the pattern of group difference on amplitude turbulence shows a tendency to reverse. Error bars represent 95% confidence intervals. Asterisks denote level of statistical significance after FDR multiple comparison correction (*** p < 0.001, ** p < 0.01, * p < 0.05).

### Variability of parcel-level functional synchronization changes as a function of well-known large-scale functional networks in autism

To better understand how variability in functional synchronization manifests in groups of parcels corresponding to well-known large-scale functional networks in autism, we utilized parcel-level turbulence metrics. Parcel-level turbulence captures the variability of synchronization of a given parcel and a given spatial scale λ across time. To investigate this, we extracted the 100 parcels with the largest absolute differences in parcel-level turbulence between autism and TD at each spatial scale. We then labeled each of these 100 parcels according to one of seven resting-state networks (RSN) identified in previous work by Yeo et al. (2011) or a subcortical network (see Figure 4a). Over short distances, the majority of parcels were those showing higher variability in functional synchronization in autism and fell within networks that deal with sensorimotor or attention processes (see Figures 4b and Supplementary Table S5). However, as spatial scale increased (e.g., decreasing λ values), the most prominent parcels were those that had decreased variability in functional synchronization in autism and were shifted towards subcortical, default mode, limbic and control networks. Note that the distribution of differences in parcel-level turbulence metrics between autism and TD followed our findings regarding amplitude turbulence (see Figure 3b and Figure 4c): at the shortest distances (e.g. λ = 0.28), the autism group exhibited higher levels of parcel-level turbulence (i.e. higher degree of variability in functional synchronization) than the TD group. However, as the distance increased (e.g. λ = 0.01), this pattern was reversed, with individual parcels of the TD group showing higher variability in functional synchronization compared to the autism group. Overall, these results suggest that highly turbulent localized dynamics are observed over sensorimotor and attention networks, while less turbulent functional dynamics over larger spatial scales occur in autism within subcortical, limbic, and other prominent networks that encompass association cortices (e.g., default mode, control networks).

**Figure 4.**
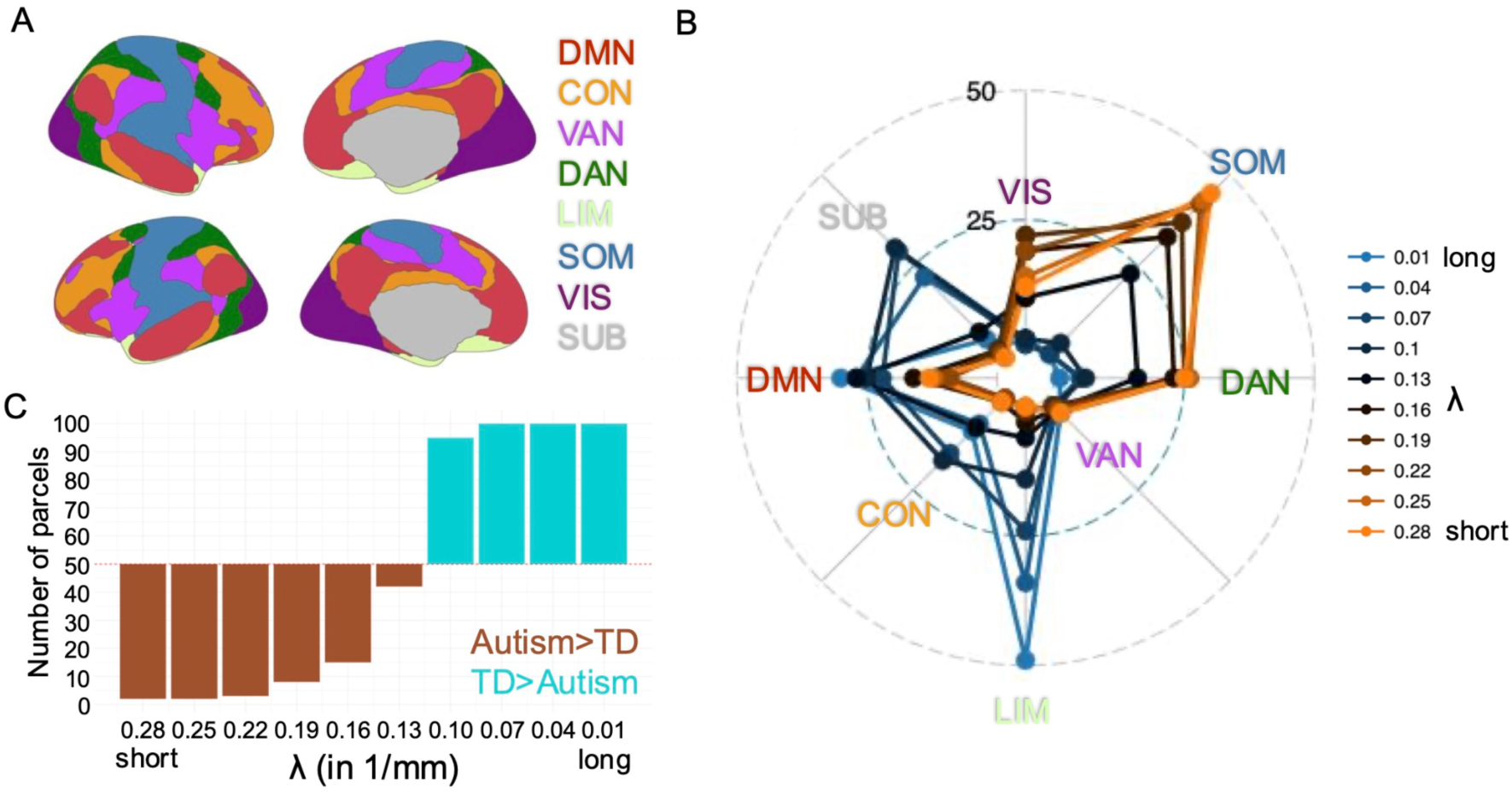
Parcel-level turbulence effects across well-known resting state networks (RSN). Panel A shows brain surface plots of 7 Yeo networks (Yeo et al., 2011) and 1 subcortical network utilized for analysis. Panel B shows spider plots that describe how the top 100 parcels with largest group differences at a given spatial scale λ were distributed across RSN networks. The different spider plots represent results at each spatial scale denoted by λ. Values within the plots indicate the percentage of parcels out of the top 100 that fall within a specific network. Panel C is a bar plot showing the percentage of parcels out of the top 100 with a TD>Autism (cyan) or Autism>TD (brown) directionality of group difference. Abbreviations: VIS, visual network; SOM, somatomotor network; DAN, dorsal-attention network; VAN, ventral-attention network; LIM, limbic network; CON, control network; DMN, default-mode network; SUB, subcortical areas.

### Variability of parcel-level functional synchronization maps onto sensorimotor-association and functional gradients in autism

The previous analysis investigated the effects of synchronization variability over well-known RSNs. RSNs represent discrete network divisions and only give one angle for examining intrinsic functional brain organization. It is known that the brain exhibits hierarchically gradient-like organization across many modalities. One hierarchical gradient we explored further is the so-called sensorimotor-association (S-A) axis (Sydnor et al., 2021) (Figure 5a). The S-A axis is hypothesized to be a neurodevelopmentally important axis for hierarchical brain organization (Sydnor et al., 2021; Tsyporin et al., 2025) and autism (Bernhardt et al., 2025) and may be one of the downstream convergent consequences of early genomic cortical patterning differences in autism (Gandal et al., 2022; Lombardo et al., 2021). The S-A axis is quite similar to primary axes of functional connectivity (FC) gradients (Margulies et al., 2016) (Figure 5b). By examining both the S-A axis and FC gradients, we provide another angle for looking at the organization of turbulence and the manifestation of group differences in autism compared to just examining RSNs. In these analyses we investigated where the top 100 regions ranked by absolute group difference would be located along the S-A and FC gradients. This analysis also sought to assess how the positioning of the top 100 regions might change as a function of spatial scale represented by the λ coefficient. To assess this change in gradient ranks as a function of λ, we used different types of models (linear, polynomial, logistic) to describe possible linear or non-linear (e.g., S-curve) relationships.

**Figure 5.**
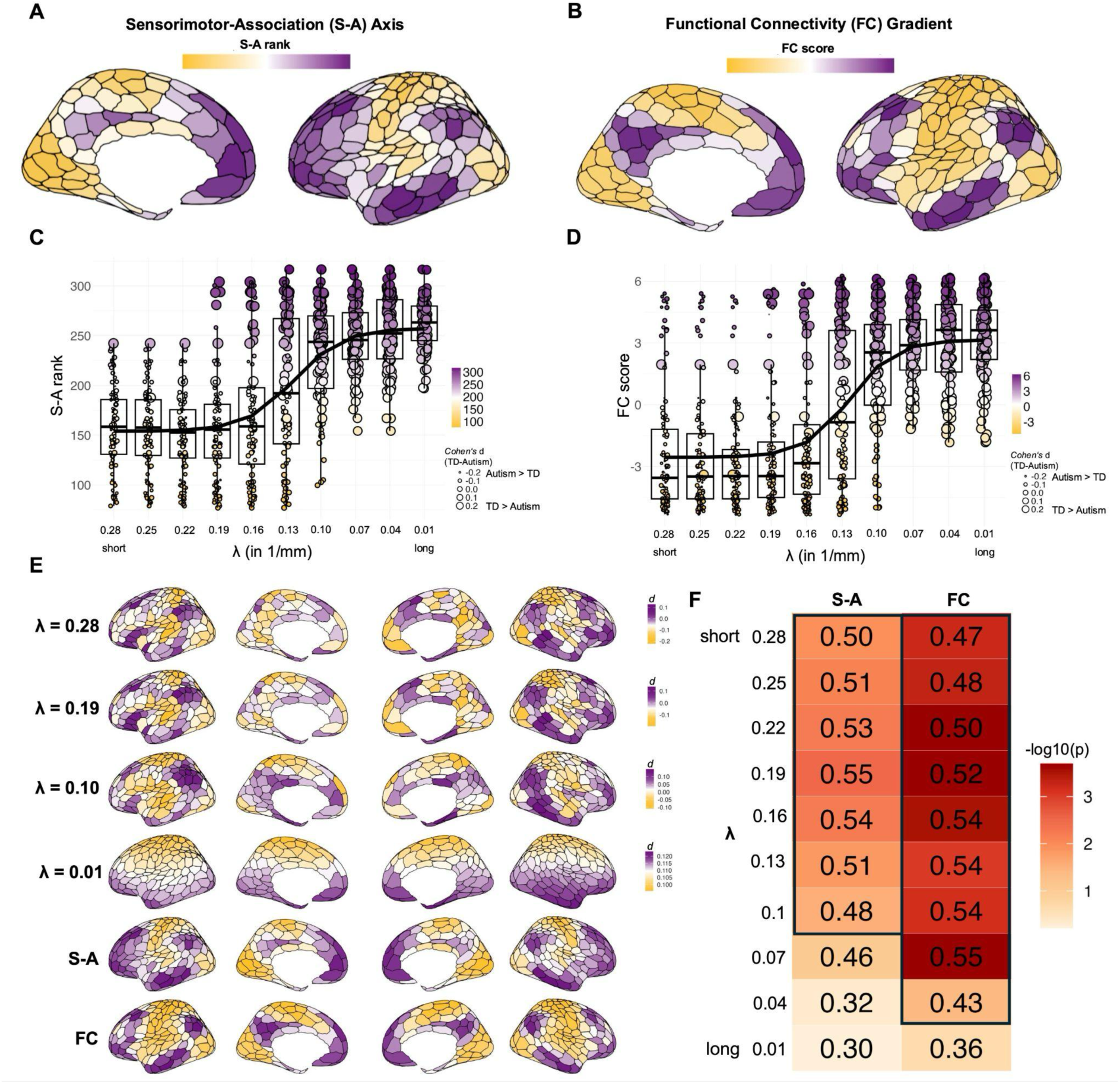
Similarity between synchronization variability group differences and S-A or functional connectivity gradient organization. Panels A and B show the S-A axis (A) and the functional connectivity gradient (B) maps. Panels C and D show ranks (shown via the y-axis and coloring of the dots) for the top 100 parcels (dots) over each spatial scale λ (x-axis). The size of the dots represents the standardized effect size for a group difference with the smallest dots representing an Autism>TD difference, while positive signed effect sizes (i.e. TD>Autism) are denoted with larger dots. The predicted values for each λ in the fitted logistic model are plotted as the smooth black curve. (E) Similarities of Cohen’s d of parcel-level turbulence and S-A and functional gradient maps. Parcel-level turbulence maps are displayed for four spatial scales, with the top panel displaying the smallest spatial scale. Golden colors indicate increased synchronization variability in autism compared to TD. Conversely, purple colors indicate decreased synchronization variability. (F) This heatmap shows the correlation values (Spearman’s ρ) of parcel-level turbulence and S-A and functional gradient maps for each spatial scale. The colors in the heatmap are -log10(p); red indicates lower p-values, and orange indicates higher p-values. For each spatial scale, the Spearman rho value is written inside the cell. Spatial scales for which there was a statistically significant correlation with S-A or FC gradients are outlined in black.

Here we find that the top 100 parcels with the largest turbulence group differences are located systematically at different ends of the S-A and FC gradients but also depend on which spatial scale is being examined. The positioning of the top 100 parcels along S-A and FC gradients changes as a function of spatial scale (i.e. λ), and that change can best be described as following an ‘S-curve’ (Figure 5c, 5d and Supplementary Table S6). This S-curve can be described by sensorimotor regions dominating the top 100 parcels at small spatial scales (i.e. higher λ), whereas association areas dominate the top 100 at large spatial scales (i.e. lower λ). Another important feature of this result is the directionality of turbulence group differences shown by the size of the dots in Figures 5c and 5d. For both S-A and FC gradients, the effect sizes are predominantly negative at small spatial scales and indicate an Autism>TD group difference. In contrast, at large spatial scales the sign of the group difference flips, indicating a TD>Autism group difference. These results suggest that at small spatial scales (i.e. higher λ), the most important regions with turbulence group differences are located at the sensorimotor pole of the S-A and FC axes and are represented by higher levels of variability in functional synchronization in autism. Conversely, at larger spatial scales (i.e. lower λ) the regions with largest group differences in turbulence are located at the association pole of S-A and FC axes and manifest as lower levels of variability in functional synchronization in autism.

In the previous analysis, we focused on the top 100 parcels showing the biggest absolute differences in parcel-level turbulence between the autism and TD groups. However, we can also statistically assess similarity in turbulence group differences and S-A or FC maps without any selection criteria (e.g., top 100 parcels). To do this, we calculated parcel-level turbulence group difference maps and then used Spearman ρ and spatial permutation spin tests to test for statistically significant similarity with the S-A or FC maps. Here we found significant correlations with S-A and FC maps at almost all spatial scales except the very large ones (e.g., λ = 0.07-0.01) (Figure 5f) (Supplementary Table S7). One possible reason for the lack of similarity at large spatial scales could be that such finer grained spatial patterning breaks down when spatial scales become too large. This can be seen in the effect size maps at λ = 0.01 in Figure 5e, as the spatial effects here are too coarse and resemble simpler dorsal-ventral gradient patterning rather than spatially more complex and fine-grained patterning consistent with S-A and FC gradients. However, overall, these findings provide further evidence that differences in synchronization variability align with well-established S-A and FC gradients across the whole cortex. Autism can be characterized by higher synchronization variability in sensorimotor areas and lower synchronization variability in association areas.

### Weaker, less spatially correlated, and rapidly decaying functional synchronization over space in autism

In addition to assessing synchronization *variability* as a function of spatial scale, our framework allowed us to also evaluate synchronization *strength*. First, we assessed synchronization strength at the whole-brain level (i.e., mean(KGOP) or *global synchronization*). We found a highly significant difference between groups (*F*(1,1003) = 22.62, *p* = 0.000002, *d* = 0.300, Figure 6a and Supplementary Table S3) with weaker whole-brain synchronization in autism compared to TD. We also investigated the strength of functional synchronization over time as a function of spatial scale (λ) - what we call *scale-specific synchronization* (i.e. mean(KLOP)). Here we find a main effect of diagnosis (*F*(1,996) = 20.90, *p* = 0.000005; Figure 6b and Supplementary Table S4) as well as a significant λ×diagnosis interaction (*F*(1,1007) = 15.23, *p* = 0.0001). Post-hoc tests revealed that scale-specific synchronization levels were significantly lower in autism than in TD for all spatial scales, with the difference being most pronounced over longer distances. These results indicate that the strength of synchronization is weaker in the autism group than in the TD group for all distances.

**Figure 6.**
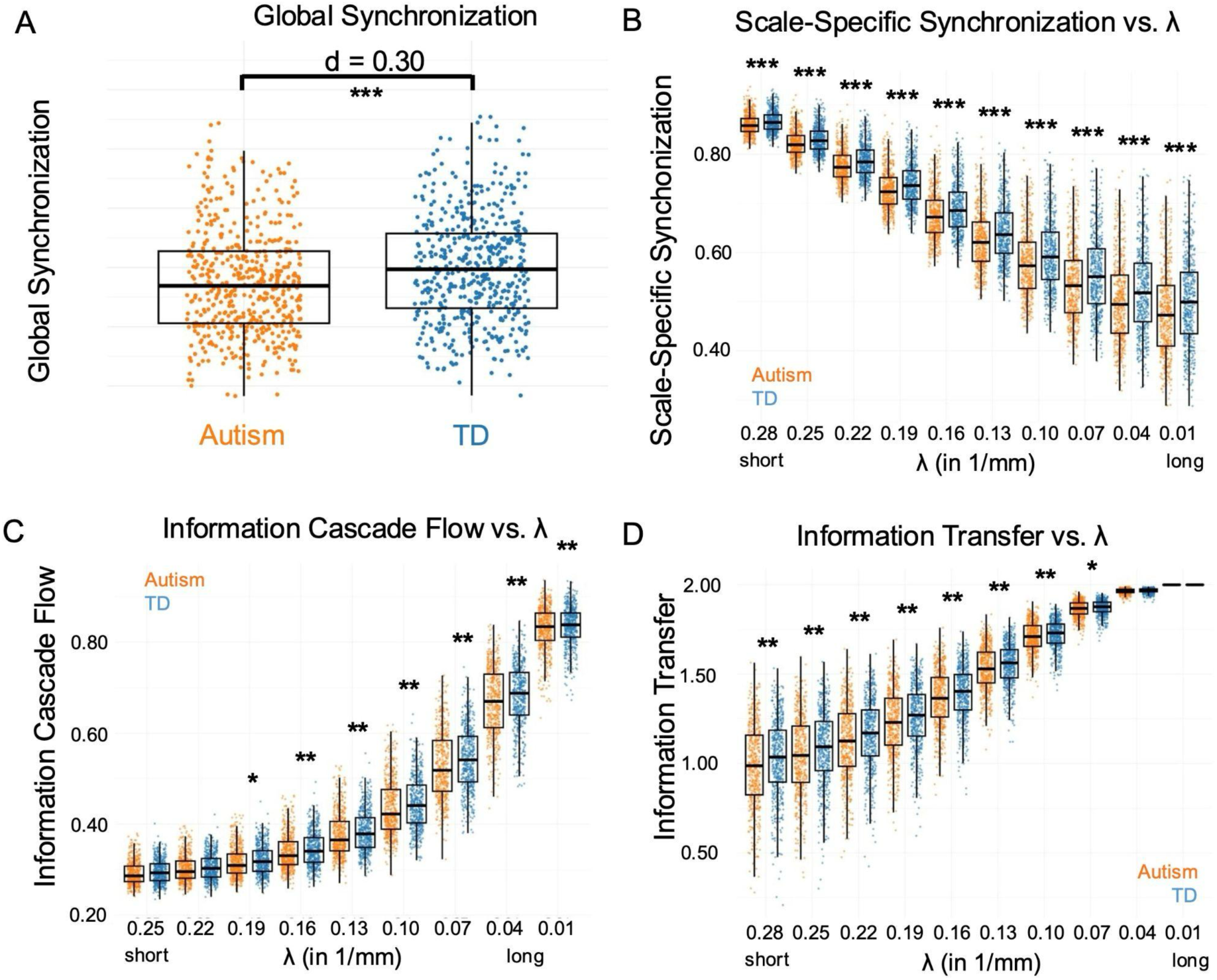
Group differences in synchronization strength, information cascade flow, and information transfer. Panel A shows a reduction in whole-brain global synchronization strength in autism (orange) relative to TD (blue). Panel B shows reduced synchronization strength in autism is also apparent at each spatial scale depicted by λ. Panels C and D depict information cascade flow (C) and information transfer levels (D) are decreased in autism relative to TD for most spatial scales (λ shown on the x-axis). Differences in information cascade flow levels are most pronounced on longer distances, while differences in information transfer levels are most pronounced on smaller distances. Asterisks denote level of statistical significance after FDR multiple comparison correction (*** p < 0.001, ** p < 0.01, * p < 0.05).

In addition to strength and variability in functional synchronization, we also were interested in testing how synchronization cascades over different spatial scales (i.e. progressively larger distances). To test this we used a measure called *information cascade flow*, or the correlation between functional synchronization at different spatial scales (e.g., corr(KLOP_λ1_, KLOP_λ2_), whereby λ1 and λ2 are adjacent spatial scales). Here we find a main effect of diagnosis (*F*(1,1020) = 9.04, *p* = 0.003) and no λ×diagnosis interaction (*F*(1,2532) = 2.12, *p* = 0.15). Follow-up tests revealed that the main effect of diagnosis is driven by lower levels of information cascade flow in autism at almost all spatial scales, with the effect being most pronounced at larger spatial scales (e.g., lower λ levels; see Figure 6c and Supplementary Table S4). This result suggests that functional synchronization across larger spatial scales is reduced in autism, likely impeding long-range information propagation.

Finally, we examined the rate of decay in functional synchronization over spatial scales, indicated by the measure we call *information transfer* (e.g., linear modeling of KLOP correlations over parcels as a function of distance). Here we find a main effect of diagnosis (*F*(1,995) = 9.07, *p* = 0.003), as well as a significant λ×diagnosis interaction (*F*(1,1073) = 10.31, *p* = 0.001). Follow-up tests on each level of λ were used to decompose the interaction effect. This revealed significantly lower levels of information transfer in autism compared to TD for all λ except for ones indicative of the largest spatial scales (λ = 0.01, λ = 0.04; see Figure 6d and Supplementary Table S4). These results suggest that functional synchronization over shorter distances decays more rapidly across space in autism compared to TD individuals. Taken together, we observed the following in autism: (1) decreased functional synchronization strength across all distances (i.e., scale-specific synchronization); (2) altered propagation of synchronization across medium and large spatial scales (i.e., information cascade flow); and (3) faster spatial decay of synchronization at short and medium distances (i.e., information transfer).

## Discussion

In this study, we applied the turbulence-dynamics framework to examine functional synchronization patterns on different spatial scales in resting-state fMRI scans of the brains of people with autism. Our findings reveal robust evidence of altered functional synchronization patterns within and between spatial scales. Compared to typically developing individuals, we observe increased variability (i.e. increased amplitude turbulence), faster spatial decay (i.e. lower information transfer), and decreased functional synchronization strength over short distances in autism. This increased variability is primarily attributed to the sensorimotor and unimodal areas of the brain. At long distances, the strength of functional synchronization remains reduced. Additionally, however, we found that the influence of lower spatial scales on adjacent, higher spatial scales of functional synchronization was reduced (i.e. information cascade flow). Furthermore, the marginally decreased variability of synchronization at long distances in individuals with autism is primarily found in the association and transmodal areas of the brain.

How do these observed results from the turbulence framework fit into long-standing ideas about altered functional connectivity as a convergent phenomenon explaining the neurobiology behind the autisms? One theme in the literature suggests that autism can be characterized by local overconnectivity versus long-range underconnectivity (Belmonte et al., 2004; Courchesne & Pierce, 2005; Hull et al., 2017). For example, using a static functional connectivity approach that measures connectivity over the entire time-series, Weber et al. (2024) found evidence supporting this view on local over-connectivity and long-range underconnectivity. However, a meta-analysis by Hull and colleagues suggest that evidence supporting this theme is mixed and may be due to methodological differences (Hull et al., 2017). As an alternative to static connectivity approaches, in this work we used a dynamic approach that enables characterization of synchronization strength, but also can allow for statements about variability in synchronization. Regarding the strength of synchronization, we find evidence that partially supports the view of underconnectivity. However, our observations show that underconnectivity is present at both local and long-range distances. The framework can additionally make observations about the variability in connectivity over time. Here we find that variability in connectivity depends on distance - shorter distances show increased variability while at longer distances group differences in variability are becoming much smaller and may change in directionality. Thus, the turbulence framework allows for a more complete description about connectivity in the autistic brain not only in terms of strength over an assumed period of time, but also about how variable connection strength is in autism. Future work using this framework in autism could help confirm whether such observations are a hallmark of autism or perhaps a subset of autistic cases with specific biological or phenotypic profiles.

### From local chaos to global decoupling: Multiscale connectivity alterations in autism

The combination of local increases in synchronization variability, faster dissipation of synchronization over space, and reduction in synchronization strength in autism may be indicative of a situation we interpret as ‘enhanced local chaos’ and may also signal something important about the underlying neurobiology residing locally at the level of synapses. Past work has shown that normative processes controlled by mTOR-dependent signalling induce synaptic spine pruning in early brain development (Lee, 2015). These processes are likely altered in autism (Pagani et al., 2021; Rosina et al., 2019) and could cause observed phenomenon in post-mortem data such as increased synaptic spine density in autism (Tang et al., 2014). These and other microstructural phenomena, such as increased cell density (Casanova et al., 2006; Courchesne et al., 2011, 2024), could help to explain the ‘enhanced local chaos’ indicated by increased synchronization variability (i.e. higher turbulence), faster dissipation of synchronization over space (i.e. information transfer), and a reduction in the strength of local functional synchronization.

In addition to a situation of ‘enhanced local chaos’, other findings from this study such as reduced information cascade flow may also hint at the idea of ‘global decoupling’ in autism. Information cascade flow can be interpreted as how synchronization travels from smaller to larger spatial scales. A reduction in this measure indicates that information at local levels does not cascade well across longer distances. This result could implicate issues with longer-range anatomical connections such as white matter tracts. Compatible with this idea, diffusion imaging studies have found reduced levels of integrity of long-range white matter tracts (i.e. reduced fractional anisotropy and diffusivity) in autism at ages like those in the current work (Aoki et al., 2013; Catani et al., 2016; Shukla et al., 2011). Thus, abnormalities in long-range white matter pathways may constrain synchronization propagation, particularly at larger spatial scales. These ideas also match well with insights from computational models. For example, Deco et al. (2021) demonstrated that long-range connections increase cascade levels in the brain. Therefore, disruption of these connections in autism may explain the reduced capacity to integrate synchronization across different spatial scales (i.e., reduced information cascade flow levels).

Taken together, these results suggest that in the brains of people with autism, local overconnectivity may give rise to excessively chaotic and turbulent synchronization dynamics at smaller spatial scales. This locally unstable pattern appears unable to sustain coherence over longer distances, resulting in reduced synchronization strength and variability. At larger scales, long-range underconnectivity and alterations in white matter anatomy may further exacerbate these disruptions, yielding a decoupled and overly rigid brain organization at longer distances. This scale-dependent dichotomy is significant as it extends previous reports of scale-independent hypervariability in functional synchronization (Mash et al., 2019).

### Altered synchronization patterns across varying distances relate to hierarchical brain organization

Moving beyond distance-dependent connectivity patterns, it has been suggested by a substantial body of research that the human brain is hierarchically organized along a gradient that spans sensorimotor to association regions (aka the S-A axis) (Margulies et al., 2016; Sydnor et al., 2021). The S-A axis has been identified as atypical in autism, as indicated by a contracted functional hierarchy observed in rsfMRI data (Hong et al., 2019). We find additional evidence implicating the S-A axis in autism with the turbulence framework applied to rsfMRI data. In particular, we observe increased short-range functional variability in sensorimotor areas alongside decreased long-range functional variability in association areas. These results may match well with observations that suggest that a diverse range of phenomena across the information processing hierarchy are different in autism (Bernhardt et al., 2025). The S–A axis is also strongly linked to cortical maturation trajectories. Markers such as the age at which peak myelination growth occurs (Baum et al., 2022) and parvalbumin expression patterns (Larsen et al., 2023) vary systematically along this axis (Sydnor et al., 2021). Thus, the differences in synchronization variability that we observed may reflect differences in the timing of cortical development along this axis. Early-developing sensorimotor regions may undergo refinement that promotes heightened local coupling, while later-developing association regions may exhibit reduced large-scale synchronization. Therefore, the turbulence alterations we observe may index developmental differences in how functional organization unfolds along the S–A axis.

### Limitations and future directions

There are some caveats and limitations relevant to this work, which can be addressed in future work. First, while we have observed robust evidence for case-control differences, most differences are small in effect size. While such small effect sizes are in keeping with many other studies examining case-control differences (Rødgaard et al., 2019), future work should apply the turbulence framework to examine important autism subtypes (Leyhausen et al., 2025; Litman et al., 2025; Lombardo et al., 2019; Mandelli et al., 2024; Rødgaard et al., 2019). Future work should also endeavour to replicate these findings in independent datasets. We have opted for maximal statistical power in the current study, utilizing all available data from the ABIDE datasets. However, future work with similarly large sample sizes would be useful to test for replicability of such effects. Finally, given the reliance on the ABIDE datasets, the current work is limited to observations on older autistic individuals, many of whom have average to high levels of intellectual ability. Future work in younger populations and over a wider range of severity and intellectual ability will be needed to generalize the results to the overall autistic population. Another key question is whether the developmental trajectories of turbulence dynamics differ between autistic and typically developing individuals. Longitudinal studies will be essential for determining how these dynamics evolve over time in each group.

In summary, our findings demonstrate that functional synchronization profiles in autism depend on spatial scale and vary systematically along the sensorimotor–association gradient. Increased local chaos, reflected in heightened turbulent dynamics, fails to propagate a stable synchronization pattern across spatial scales. As a result, autistic individuals show a decoupled and overly rigid synchronization profile over longer cortical distances. When projected onto the sensorimotor–association axis, these effects manifest as increased synchronization variability within sensorimotor regions but reduced variability within association cortices. As next steps, more research is needed to replicate these findings and to further explore differences in the turbulence framework outcomes across clinically relevant autism subtypes.

## Data Availability

All data produced are available online at: https://fcon_1000.projects.nitrc.org/indi/abide/

## Acknowledgments

This work was supported by funding from the European Union’s Horizon Europe research and innovation programme under grant agreement No 101087263 (AUTISMS-3D) (ERC Consolidator Grant to MVL).

## Author Contributions

Conceptualization: BF, HGR, MVL. Methodology: BF, YSP, GD, MK, HGR, MVL. Formal analysis: BF, IS, MVL. Investigation: BF, IS, MVL. Writing - original draft preparation: BF, IS, MK, GD, HGR, MVL. Writing - review and editing: BF, GD, MK, HGR, MVL. Visualization: BF, MVL. Supervision: HGR, MVL. Project administration: MVL. Funding acquisition: MVL.

## Supplementary Figures

**Figure S1.**
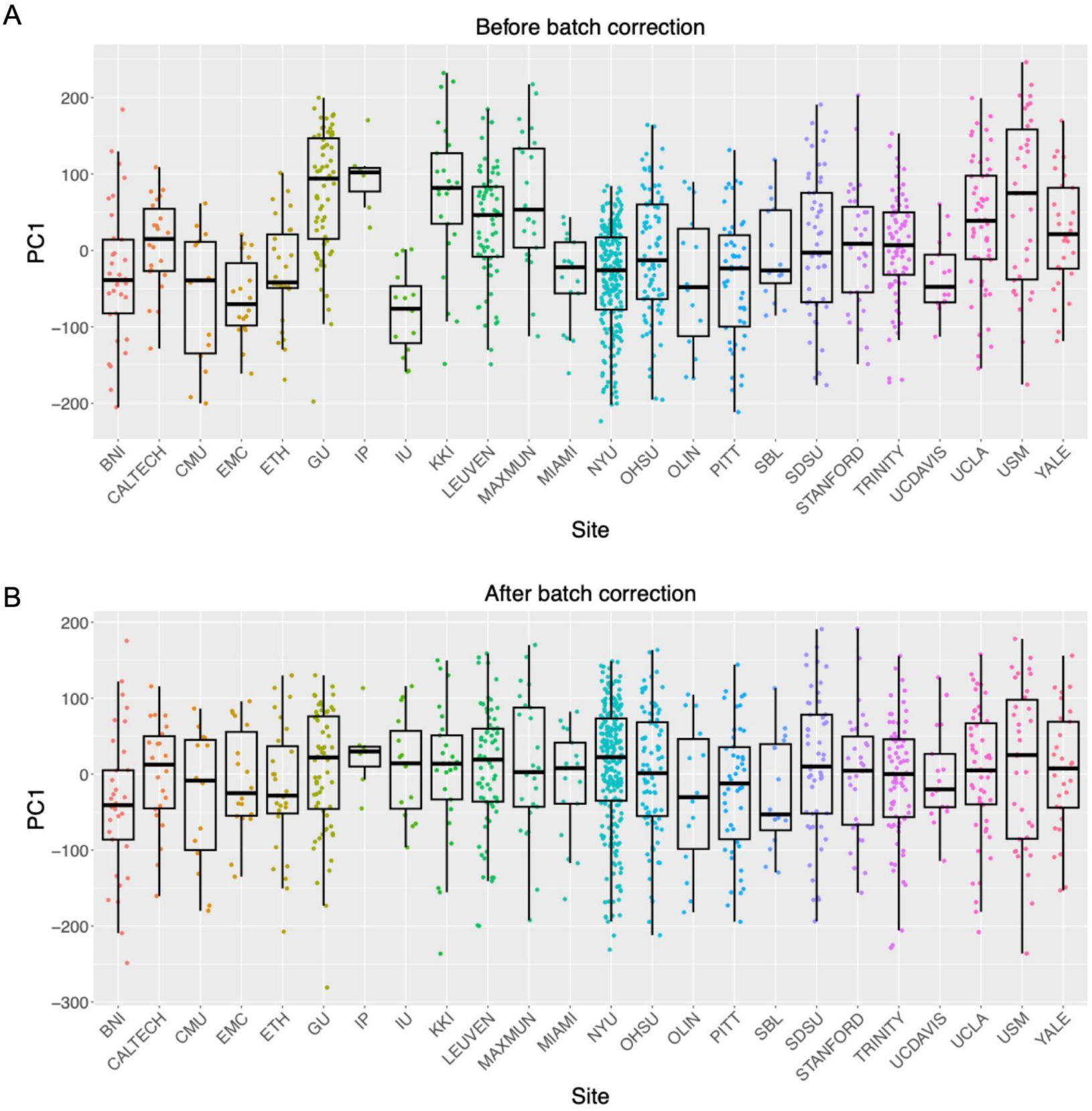
Boxplots of the first principal component values of all combined information processing measures for each site before (A) and after (B) batch correction.

## Supplementary Table Legends

***Supplementary Table 1:*** *Sample size and descriptive statistics for the ABIDE dataset. Age is stated in years*.

***Supplementary Table 2:*** *MRI acquisition parameters for different sites in the ABIDE dataset*.

***Supplementary Table 3:*** *Inferential statistics for global synchronization and metastability*.

***SupplementaryTable 4:*** *Inferential statistics for scale-specific synchronization, amplitude turbulence, information cascade flow, and information transfer measures*.

***Supplementary Table 5:*** *Parcel-level turbulence differences measured in effect sizes (Cohen’s d)*

***Supplementary Table 6:*** *Model comparisons for S-A and functional connectivity gradient, along with best model (logistic) statistics*.

***Supplementary Table 7:*** *Correlation between parcel-level turbulence difference maps (TD-autism) and S-A or FC gradients (Schaefer400 parcellation)*.

## Supplementary Movie legend

***Supplementary Movie 1:*** *This movie shows the spatiotemporal evolution of the phases and the Kuramoto Local Order Parameter (KLOP) for each of the 1,054 parcels. In the middle panel, each dot represents a single parcel. The y-value of each parcel corresponds to its phase value at a given timepoint. The brain rendering in the corner of the movie shows how this phase-information translates into the KLOP for each parcel at a given timepoint. Brighter colours represent high synchronisation (high KLOP) for a given parcel, whereas darker colours represent low synchronisation (low KLOP). The panels in the top row show a focal spatial kernel (λ = 0.28), producing a finer, more mosaic-like spatiotemporal evolution of the KLOP. Conversely, the bottom panels show a broader spatial kernel (λ = 0.01), displaying a nearly brain-wide spatiotemporal evolution of the KLOP*.

